# Establishing a core set of open science practices in biomedicine: a modified Delphi study

**DOI:** 10.1101/2022.06.27.22276964

**Authors:** Kelly D. Cobey, Stefanie Haustein, Jamie Brehaut, Ulrich Dirnagl, Delwen L. Franzen, Lars G. Hemkens, Justin Presseau, Nico Riedel, Daniel Strech, Juan Pablo Alperin, Rodrigo Costas, Emily S Sena, Thed van Leeuwen, Clare L. Ardern, Isabel O. L. Bacellar, Nancy Camack, Marcos Britto Correa, Roberto Buccione, Maximiliano Sergio Cenci, Dean A. Fergusson, Cassandra Gould van Praag, Michael M. Hoffman, Renata Moraes Bielemann, Ugo Moschini, Mauro Paschetta, Valentina Pasquale, Valeria E. Rac, Dylan Roskams-Edris, Hermann M. Schatzl, Jo Anne Stratton, David Moher

## Abstract

**Background:** Mandates and recommendations related to embedding open science practices within the research lifecycle are increasingly common. Few stakeholders, however, are monitoring compliance to their mandates or recommendations. It is necessary to monitor the current state of open science to track changes over time and to identify areas to create interventions to drive improvements.

Monitoring open science practices requires that they are defined and operationalized. Involving the biomedical community, we sought to reach consensus on a core set of open science practices to monitor at biomedical research institutions.

**Methods and Findings:** To establish consensus in a structured and systematic fashion, we conducted a modified 3-round Delphi study. Participants in Round 1 were 80 individuals from 20 biomedical research institutions that exhibit interest in or actively support open science. Participants were research administrators, researchers, specialists in dedicated open science roles, and librarians. In Rounds 1 and 2, participants completed an online survey evaluating a set of potential open science practices that could be important and meaningful to monitor in an automated institutional open science dashboard. Participants voted on the inclusion of each item and provided a rationale for their choice. We defined consensus as 80% agreement. Between rounds, participants received aggregated voting scores for each item and anonymized comments from all participants, and were asked to re-vote on items that did not reach consensus. For Round 3, we hosted two half-day virtual meetings with 21 and 17 participants respectively to discuss and vote on all items that had not reached consensus after Round 2. Ultimately, participants reached consensus to include a 19 open science practices.

**Conclusions:** A group of international stakeholders used a modified Delphi process to agree upon open science practices to monitor in a proposed open science dashboard for biomedical institutions. The core set of 19 open science practices identified by participants will form the foundation for institutional dashboards that display compliance with open science practices. They will now be assessed and tested for automatic inclusion in terms of technical feasibility. Using user-centered design, participating institutions will be involved in creating a dashboard prototype, which can then be implemented to monitor rates of open science practices at biomedical institutions. Our methods and approach may also transfer to other research settings–other disciplines could consider using our consensus list as a starting point for agreement upon a discipline-specific set of open science practices to monitor. The findings may also be of broader value to the development of policy, education, and interventions.

## Introduction

In November 2021, UNESCO adopted its Recommendation on Open Science, defining open science “as an inclusive construct that combines various movements and practices aiming to make multilingual scientific knowledge openly available, accessible and reusable for everyone, to increase scientific collaborations and sharing of information for the benefits of science and society, and to open the processes of scientific knowledge creation, evaluation and communication to societal actors beyond the traditional scientific community.”^1^ UNESCO recommends that its 193 member states take action towards achieving open science globally. The recommendation emphasizes the importance of monitoring policies and practices in achieving this goal^1^. Open science provides a means to improve the quality and reproducibility of research^2, 3^, and a mechanism to foster innovation and discovery^4, 5^. The UNESCO Recommendation has cemented open science’s position as a global science policy priority. It follows other initiatives from major research funders such as the Open Research Funders Group as well as national efforts to implement open science via federal open science plans^6, 7^.

Despite these commitments from policy makers and funders, adopting and implementing open science has not been straightforward. There remains an intense debate about how to motivate and incentivize individual researchers to adopt open science practices^8–10^, and how to best track open science practices within the community. A key concern is the need for funding to cover the additional fees and time costs needed to adhere to some open science best practices, when the academic reward system and career advancement still depend on traditional, closed research practices. What ‘counts’ in the tenure process is typically the outwardly observable number of publications in prestigious–typically high impact factor and often paywalled–journals, rather than efforts towards making research more accessible, shareable, transparent and reusable.

Monitoring open science practices is essential if the research community intends to evaluate the impact of policies and other interventions to drive improvements, and even just understanding the current adoption of open science practices in a research community. To improve their open science practices, institutions need measure their current performance, however there is presently no effective system for efficient and large-scale monitoring without significant effort.

Consider the example of open access publishing. A researcher-led large analysis of researchers’ compliance with funder mandates for open access publishing showed that the rate of adherence varied considerably by funder^18^. In Canada, the Canadian Institutes of Health Research (CIHR) had an open access requirement for depositing articles between 2008 and 2015. This deposit requirement was modified when CIHR and the other two major Canadian funding agencies harmonized their policies. The result was a drop in openly available CIHR-funded research from approximately 60% in 2014 to approximately 40% in 2017^11^. In the absence of monitoring, it is not possible to evaluate the impact of introducing a new policy or to measure how other changes in the scholarly landscape impact open science practices.

The current project aimed to establish a core set of open science practices within biomedicine to implement and monitor at the institutional level. Specifically, to standardize and effectively monitor open science we sought to agree upon a core set of practices within the biomedical research community. Our vision to establish a core set of open science practices stems from the work of the Core Outcome Measures in Effectiveness Trials (COMET)^12^. If trialists agree on a few core outcomes to assess across trials, it strengthens the totality of evidence, enables more meaningful use in systematic reviews, promotes meta-research, and may subsequently reduce waste in research. We sought to apply this concept of community agreed standardization to open science in biomedical research, which currently lacks consensus on best practices and work to operationalize different open science practices. There likely exist discipline-specific open science practices and norms, so we focused our work on biomedical research. The COVID-19 pandemic has created increased impetus for, and attention to, open science which has contributed to the development of new discipline specific practices for openess^13–15^.

The core set of open science practices identified from this work may serve the community in many ways, including developing policy, education, or other interventions to support the implementation of these practices. Most immediately, the practices can inform the development of an automated open science dashboard that can be deployed by biomedical institutions to efficiently monitor adoption of (and provide feedback on) these practices. By establishing what should be reported in an institutional open science dashboard through a consensus building process with relevant stakeholders, we aim to ensure the tool is appropriate to the needs of the community.

## Methods

### Transparency Statement

This study received ethical approval from the Ottawa Health Science Network Research Ethics Board (20210515-01H). The study protocol was posted on the Open Science Framework^16^ prior to data collection and has drawn some of the text of this Methods section directly from the protocol. All related study materials and data are available at: 10.17605/OSF.IO/JM8WG^17^.

### Study Design

We conducted a 3-round modified Delphi survey study. Delphi studies structure communication between participants to establish consensus^18^. Typically, Delphi studies use several rounds of surveys in which participants, experts in the topic area, vote on specific issues. Between rounds, votes are aggregated and anonymized and then presented back to participants along with their own individual scores, and feedback on others’ anonymized voting decisions^19, 20^. This gives participants the opportunity to consider the group’s thoughts and to compare and adjust their own assessment in the next round. A strength of this method of communication is that it allows all individuals in a group to communicate their views. Anonymous voting also limits direct confrontation among individuals and the influence of power dynamics and hierarchies on the group’s decision.

The first two rounds of the Delphi were online surveys administered using Surveylet. Surveylet is a purpose-built platform for developing and administering Delphi surveys^21^. Round 3 took the form of two half-day meetings hosted on Zoom^22^. Hosting Round 3 in the form of an online meeting is a modification of the traditional Delphi approach. This was done to provide an opportunity for more nuanced discussion among participants about the potential open science practices while still retaining anonymized online voting. We opted for a virtual meeting given the COVID-19 pandemic restrictions at the time and the cost effectiveness for enabling international participation.

### Recruitment of Participants

We generated an initial list of 32 research institutions from our networks who we felt were directly or indirectly interested in monitoring open science practices in biomedicine. We then snowball sampled (i.e., an iterative process of selecting institutions; the process is often started with a small number of institutions that meet specific inclusion criteria) from this list by asking leaders at interested institutions to connect us with their peers at other institutions who may be interested in participating. We explicitly asked institutional leaders we contacted to consider geographic representation in their suggestions.

The snowball sampling approach generated a list of 32 institutions from 22 countries and institutional contacts in leadership, or contacts who could communicate our invitation to leadership (Appendix 1). We contacted each institution to describe our study and invite them to participate. If an institution confirmed interest in participating, we asked them to identify 4–6 members of their research community from any/ all of the following groups to take part in the Delphi:

1. Library or scholarly communication staff (e.g., responsible for purchasing journal content, responsible for facilitating data sharing or management)
2. Research administrators or leaders (e.g., head of department, CEO, senior management)
3. Staff involved in researcher assessment (e.g., appointment and tenure committee members)
4. Individuals involved in institutional metrics assessment or reporting (e.g., performance management roles)

Because titles and roles differ from institution to institution, we left it to the discretion of the institution to identify participants. Broadly, we aimed to include people who either knew about scholarly metrics, or who made decisions regarding researcher assessment or hiring. We also explicitly encouraged the institutions to consider diversity of their representing participants (including gender, race) when inviting people to contribute. Institutions were sent reminders to respond after 1 and 2 weeks, respectively to confirm their interest and to have their representative participants complete the initial survey. To acknowledge participants’ time for taking part in the study, they had the opportunity to enter their name into a draw to win an iPad.

#### Round 1

Participants were asked to complete an 8-item survey via Surveylet (Appendix 2). This survey was pilot tested for useability, and we estimated a survey completion time of 15 min. The survey gathered participant demographics, information about their institution and their role within it.

Then, the survey presented participants with a list of 17 open science practices potentially relevant to include in an automated dashboard for biomedical research institutions. This list was developed by the project team. Each open science practice was defined to ensure clarity and that participants had standardized information. Participants were asked to indicate the extent to which they agreed with the inclusion of each practice in a list of core open science practices tracked in the proposed institutional dashboard. We stressed that we sought to include only practices of broad applicability and high priority in the implementation of open science in biomedicine. We instructed participants not to consider the technical feasibility of automating the presented open science practices for the proposed dashboard. Participants responded on a 9-point Likert scale with endpoints labeled ‘strongly disagree’ (1) to ‘strongly agree’ (9). Participants were also provided with a textbox after each item to provide any additional information or to justify their answer. The last part of the survey invited participants to describe additional open science practices which they believed should be considered core that were not already presented and to offer any additional feedback on the survey.

Scores for each of the survey items on Round 1 were aggregated and analyzed. All items in which 80% of respondents felt the practice should be included or excluded were removed from voting in round 2. We defined consensus as 80% of responses being in the top third (7-9) or bottom third (1-3) of the 9-point scale.

#### Round 2

For the complete Round 2 survey please see Appendix 3. All participants who completed Round 1 of the Delphi were re-invited to participate in Round 2. Any items that did not reach consensus in Round 1 were presented again in Round 2. Participants were first presented with the practice under consideration, the aggregate group score for that practice from Round 1, their own score for that practice from Round 1, and comments provided by participants. They were then asked to indicate if the practice should be: (i) ‘included in the dashboard’; (ii) ‘excluded from the dashboard’; (iii) ‘discussed at the consensus meeting’, or to indicate that they (iv) ‘don’t have expertise related to the topic’. Participants were also provided with all new open science practices suggested by participants in Round 1. In total 10 new practices were presented in Round 2. Each potential new item was briefly defined to ensure clarity and standardized information about the practice. As in Round 1, it was stressed that the dashboard sought to include only open science practices that were essential to implementing open science in biomedicine. Responses to the new items were provided on the same 9-point Likert scale used in survey 1, with endpoints labeled ‘strongly disagree’ (1) to ‘strongly agree’ (9). Again, participants could provide additional information or justify their answer. At the end of the survey, participants could again suggest new items to include, and provide any additional feedback on the survey.

#### Round 3

For the list of items voted on in round 3, please see Appendix 4. We randomly selected 50 of the 80 respondents from Round 1 to invite to participate in Round 3 (using the RAND function in Excel). Given that Round 3 required virtual attendance at two half-day meetings, we anticipated approximately half of those invited would participate which would provide a feasible group size for online discussion. We made slight modifications to our list of randomly selected invitees to ensure all institutions were sufficiently represented. Specifically, we added a randomly selected participant from any institution not represented in the list and when doing so removed a random participant from the institution with highest representation. Due to time-zone scheduling difficulties we replaced all participants from Australia. The materials used for the meeting, including the agenda, have been shared on the Open Science Framework.

To minimize the potential for bias in the discussion introduced by the core research team, the online meeting was moderated by a researcher (CLA) experienced with Delphi studies and familiar with research assessment, but who was not part of the initial core research team. On day 1 we presented the results of Rounds 1 and 2 of the Delphi survey, and highlighted items that reached consensus. We also presented a prototype of what the dashboard tool could look like (https://quest-dashboard.charite.de/). Then, we gave participants the opportunity to engage in a general discussion about the project. Following this, the items not yet in consensus were grouped according to themes and presented for discussion and voting for inclusion/exclusion in the dashboard. Participants voted on each of the items using Zoom polling software. Responses were recorded using the following options: (i) “include”, (ii) “exclude”, or (iii) “abstain”. In accordance with Rounds 1 and 2, consensus was prospectively defined as 80% of the responses in the include or exclude category, after removal of “abstain” responses. The wording of some items was modified based on discussion to better reflect nuances in the views of the group prior to voting.

After voting was completed on all potential open science practices, we facilitated a focused discussion on how to best implement the proposed open science dashboard. An expert (JB) in implementing complex interventions based on and French’s Framework^29^ for theory-informed behaviour change interventions provided a presentation to offer context. This was followed by a brainstorming exercise structured around addressing the four steps of French’s Framework in regards to implementing the open science dashboard: Step 1) Who needs to do what, differently? ; Step 2) Which barriers and enablers need to be addressed?; Step 3) Which intervention components could overcome the modifiable barriers and enhance the enablers?; Step 4) How can behaviour changes be measured and understood? Following the second day of the meeting, participants were sent the list of the core open science practices established through the Delphi process and asked to rank order the relative importance of each for prioritizing in the proposed dashboard. Based on discussions on day 2 of the meeting, items were split into two categories, ‘traditional open science practices’ and broader ‘transparency practices’. See Appendix 5.

### Protocol Amendments

We had indicated that we would post information about our study, to recruit new institutions, to the Declaration on Research Assessment (DORA) newsletter. This was not pursued as we felt our snowball sampling from our originally generated list was sufficient. Sticking to our snowballing approach also allowed us to retain a measure of the population size of institutions invited and the response rate.

## Results

### Round 1

#### Participants

We excluded participants who did not complete 80% or more of the survey in this round. A total of 80 participants from 20 institutions in 13 countries completed Round 1. Full demographics are described in Table 1. A total of 44 (55.0%) of participants identified as men, 35 (43.8%) as women, and 1 (1.3%) as another gender. Of the 32 research institutions that were identified to contribute to the study, 20 (62.5%) ended up contributing, and 1-7 participants from each organization responded to our survey. Researchers (N= 31, 38.8%) and Research Administrators (N=18, 22.5%) comprised most of the sample.

**Table 1.**
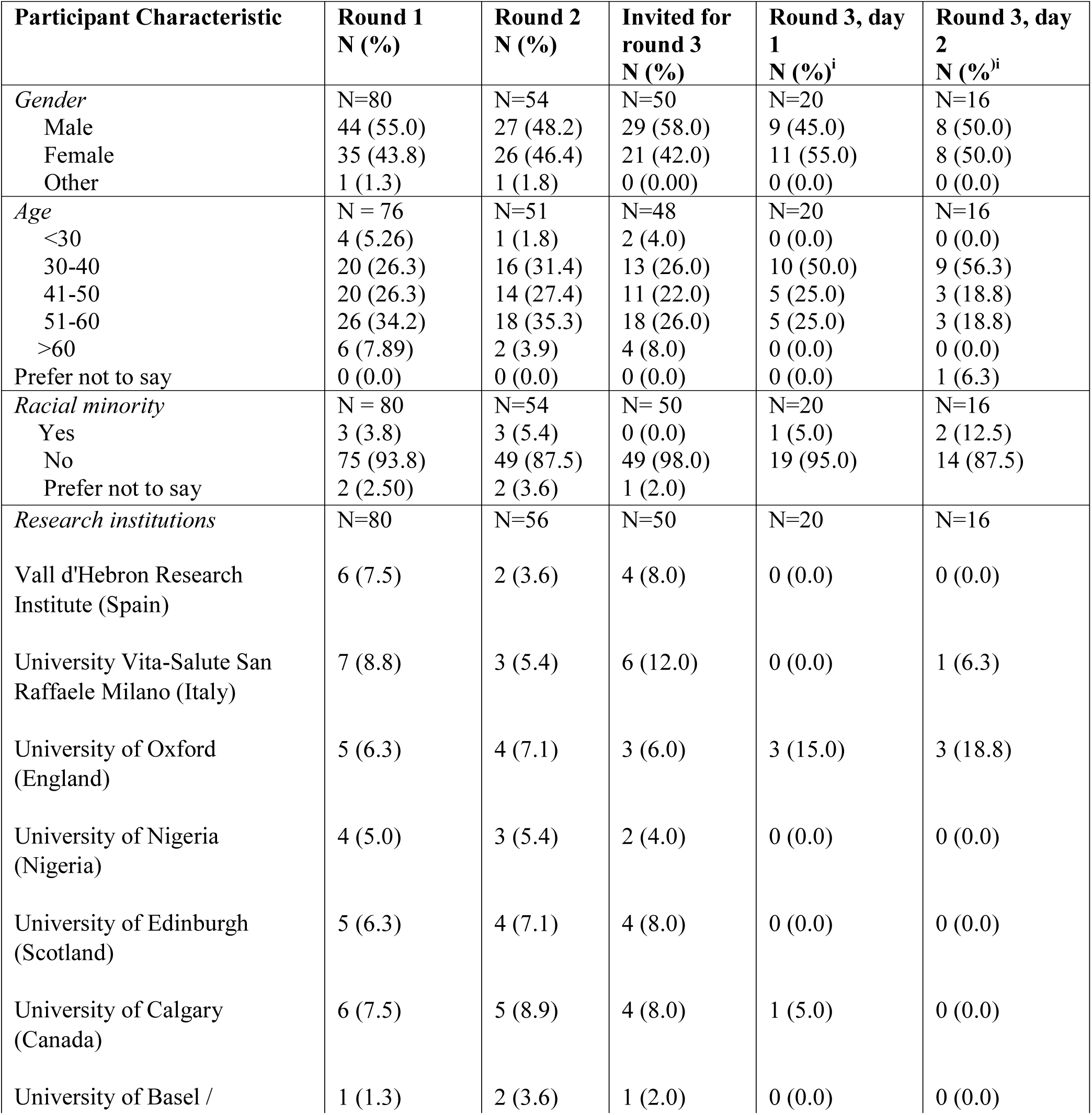
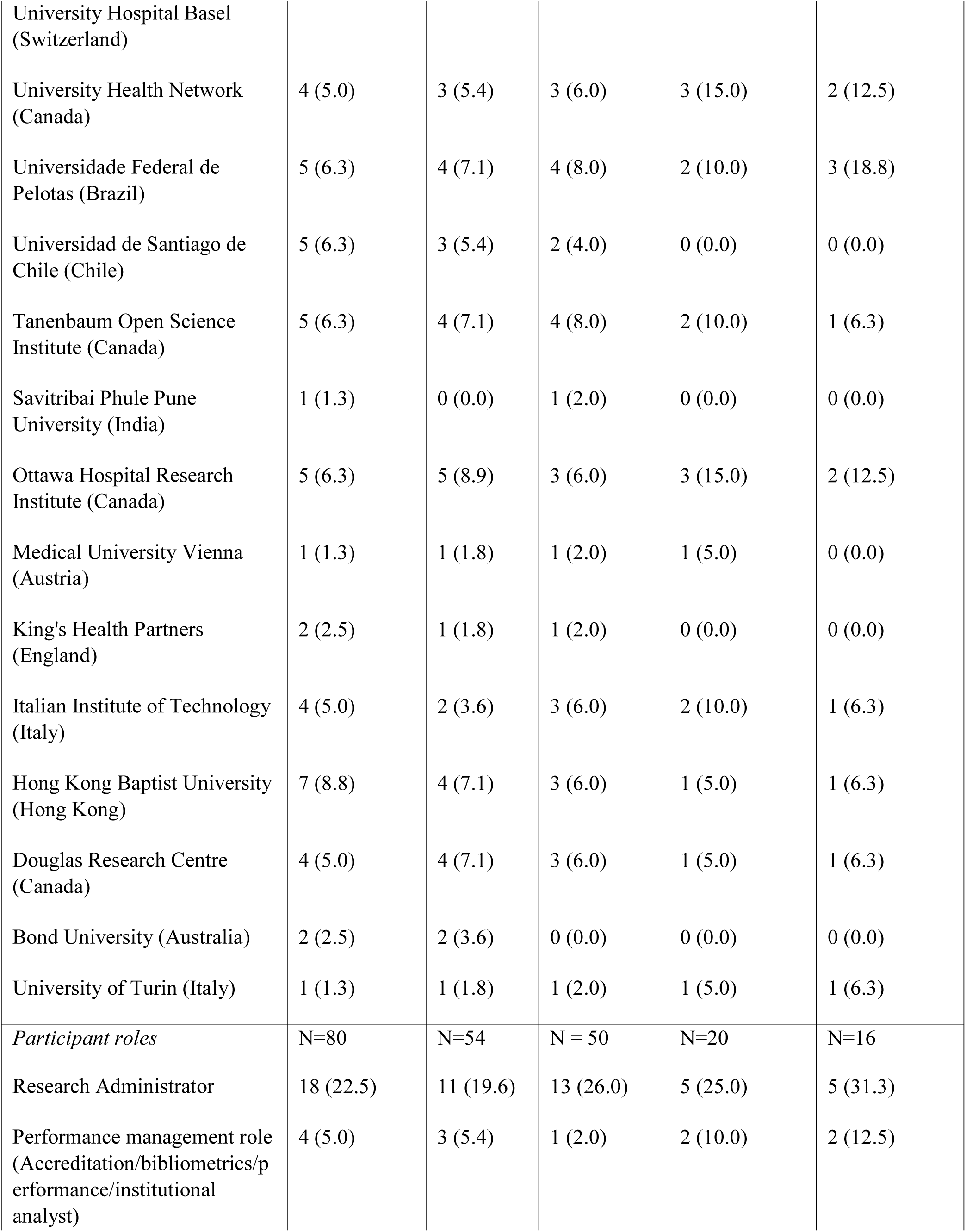
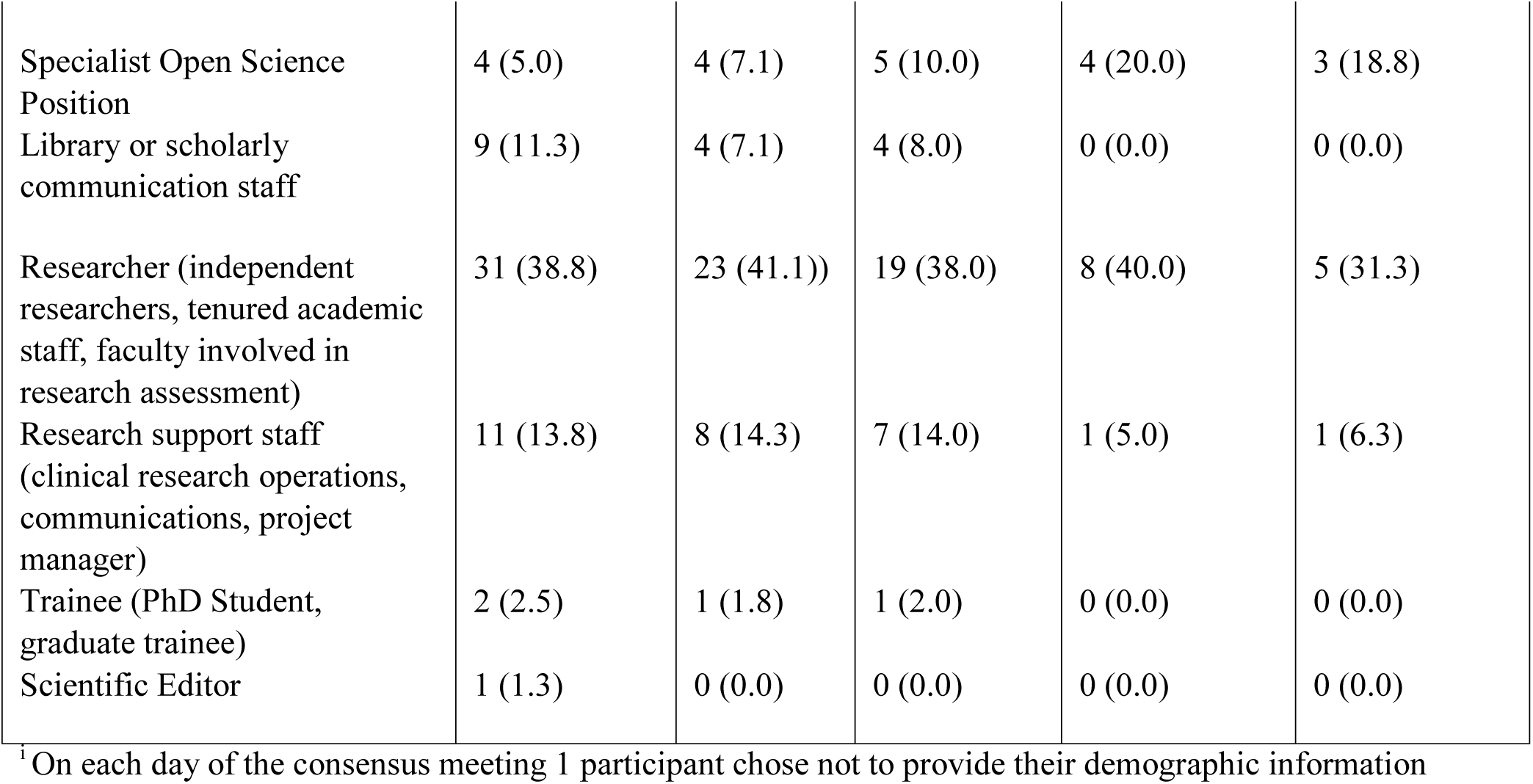
Participant Characteristics

#### Round 1 voting

Of the 17 potential core open science practices presented in Round 1, two reached consensus. Participants agreed that 1) registering clinical trials on a registry prior to recruitment, and 2) reporting author conflicts of interest in published articles were essential to include. See full results in Table 2.

**Table 2.** Delphi voting results by round. Bolded numbers indicate consensus.

|  |  | Round 1 |  | Round 2 |  | Round 3 |  |
| --- | --- | --- | --- | --- | --- | --- | --- |
| # | Open Science Practice | Scale points | N (%) | Scale | N (%) | Scale | N (%) |
| 1 | whether clinical trials have been registered before starting recruitment | 1-3<br>4-6<br>7-9 | 9 (11.7)<br>4 (5.2)<br>**64 (83.1)** |  |  |  |  |
| 2 | whether systematic reviews have been registered before data collection | 1-3<br>4-6<br>7-9 | 6 (7.9)<br>14 (18.4)<br>56 (73.7) | Include<br>Exclude<br>Discuss<br>No expertise | 36 (75.0)<br>7 (14.6)<br>5 (10.4)<br>10 | Include<br>Exclude<br>Abstain | **15 (88.2)**<br>2 (11.8)<br>4 |
| 3 | whether hypothesis testing research has been registered | 1-3<br>4-6<br>7-9 | 17 (22.1)<br>18 (23.4)<br>42 (54.5) | Include<br>Exclude<br>Discuss<br>No expertise | 26 (51.0)<br>18 (35.3)<br>7 (13.7)<br>5 | Include<br>Exclude<br>Abstain | 7 (46.7)<br>8 (53.3)<br>5 |
| 4 | whether any research paper has been registered | 1-3<br>4-6<br>7-9 | 13 (16.7)<br>28 (35.9)<br>37 (47.4) | Include<br>Exclude<br>Discuss<br>No expertise | 15 (28.8)<br>28 (53.8)<br>9 (17.3)<br>4 | Include<br>Exclude<br>Abstain | 7 (41.2)<br>10 (58.8)<br>3 |
| 5 | whether data was shared openly at the time of publication (with limited exceptions) | 1-3<br>4-6<br>7-9 | 11 (14.1)<br>13 (16.7)<br>54 (69.2) | Include<br>Exclude<br>Discuss<br>No expertise | **46 (85.2)**<br>3 (5.6)<br>5 (9.3)<br>3 |  |  |
| 6 | whether code was shared openly at the time of publication (with limited exceptions) | 1-3<br>4-6<br>7-9 | 4 (5.1)<br>18 (23.1)<br>56 (71.8) | Include<br>Exclude<br>Discuss<br>No expertise | **46 (86.8)**<br>4 (7.5)<br>3 (5.7)<br>4 |  |  |
| 7 | whether study materials were shared openly at the time of publication (with limited exceptions) | 1-3<br>4-6<br>7-9 | 11 (14.1)<br>17 (21.8)<br>50 (64.1) | Include<br>Exclude<br>Discuss<br>No expertise | 34 (70.8)<br>6 (12.5)<br>8 (16.7)<br>7 | Item modified for voting<br>(See #8 below) |  |
| 8 | whether there was a statement about study materials sharing with publications |  |  |  |  | Include<br>Exclude<br>Abstain | **18 (94.7)**<br>1 (5.3)<br>2 |
| 9 | reporting the number of preprints | 1-3<br>4-6<br>7-9 | 13 (16.9)<br>21 (27.3)<br>43 (55.8) | Include<br>Exclude<br>Discuss<br>No expertise | 30 (57.7)<br>13 (25.0)<br>9 (17.3)<br>4 | Include<br>Exclude<br>Abstain | **15 (88.2)**<br>2 (11.8)<br>0 |
| 10 | reporting whether articles were published Open Access at the time of publication | 1-3<br>4-6<br>7-9 | 10 (13.0)<br>14 (18.2)<br>53 (68.8) | Include<br>Exclude<br>Discuss<br>No expertise | 34 (64.2)<br>8 (15.1)<br>11 (20.8)<br>3 | Item modified for voting<br>(See #12 below) |  |
| 11 | reporting whether articles were published Open Access but allowing an embargo period (e.g., 1 year) | 1-3<br>4-6<br>7-9 | 23 (30.3)<br>27 (35.5)<br>26 (34.2) | Include<br>Exclude<br>Discuss<br>No expertise | 28 (53.8)<br>14 (26.9)<br>10 (19.2)<br>4 | Item modified for voting<br>(See #12 below) |  |
| 12 | reporting what proportion of articles are published open access with a breakdown of time delay |  |  |  |  | Include<br>Exclude<br>Abstain | **20 (100)**<br>0 (0)<br>1 |
| 13 | reporting whether reporting guideline checklists were used | 1-3<br>4-6<br>7-9 | 5 (6.4)<br>13 (16.7)<br>60 (76.9) | Include<br>Exclude<br>Discuss<br>No expertise | **39**<br>**(83.0)**<br>4 (8.5)<br>4 (8.5)<br>10 |  |  |
| 14 | reporting whether author conflicts of interests were declared | 1-3<br>4-6<br>7-9 | 9 (11.5)<br>4 (5.1)<br>**65 (83.3)** |  |  |  |  |
| 15 | reporting whether author contributions were described | 1-3<br>4-6<br>7-9 | 5 (6.4)<br>17 (21.8)<br>56 (71.8) | Include<br>Exclude<br>Discuss<br>No expertise | **44**<br>**(80.0)**<br>6 (10.9)<br>5 (9.1)<br>1 |  |  |
| 16 | reporting on the use of open lab notebooks | 1-3<br>4-6<br>7-9 | 18 (23.1)<br>32 (41.0)<br>28 (35.9) | Include<br>Exclude<br>Discuss<br>No expertise | 13<br>(28.9)<br>26<br>(57.8)<br>6 (13.3)<br>12 | Include<br>Exclude<br>Abstain | 6 (35.3)<br>11 (64.7)<br>4 |
| 17 | reporting whether ORCID identifiers were used | 1-3<br>4-6<br>7-9 | 9 (11.5)<br>11 (14.1)<br>58 (74.4) | Include<br>Exclude<br>Discuss<br>No expertise | **48**<br>**(87.3)**<br>5 (9.1)<br>2 (3.6)<br>2 |  |  |
| 18 | reporting that registered clinical trials were reported in the registry within 2 years of study completion | 1-3<br>4-6<br>7-9 | 9 (11.5)<br>11 (14.1)<br>58 (74.4) | Include<br>Exclude<br>Discuss<br>No expertise | **45**<br>**(93.8)**<br>1 (2.1)<br>2 (4.2)<br>8 | Item re-voted on with 1 year timeframe (See item #19 below) |  |
| 19 | reporting that registered clinical trials were reported in the registry within 1 year of study completion |  |  |  |  | Include<br>Exclude<br>Abstain | **11 (91.7)**<br>1 (8.3)<br>5 |
| 20 | reporting the number of replication studies | 1-3<br>4-6<br>7-9 | 11 (14.1)<br>22 (28.2)<br>45 (57.7) | Include<br>Exclude<br>Discuss<br>No expertise | 28 (58.3)<br>14 (29.2)<br>6 (12.5)<br>7 | Item modified for voting<br>(See items #21 and #22 below) |  |
| 21 | reporting citations to data |  |  |  |  | Include<br>Exclude<br>Abstain | **17 (89.5)**<br>2 (10.5)<br>3 |
| 22 | reporting citations to code |  |  |  |  | Include<br>Exclude<br>Abstain | 14 (77.8)<br>4 (22.2)<br>2 |
| 23 | reporting on the use of Research Resource Identifiers (RRID) (where relevant) |  |  | 1-3<br>4-6<br>7-9 | 9 (17.3)<br>25 (48.1)<br>18 (34.6) | Include<br>Exclude<br>Abstain | 7 (53.8)<br>6 (46.2)<br>4 |
| 24 | reporting whether research articles include funding statements |  |  | 1-3<br>4-6<br>7-9 | 8 (15.4)<br>7 (13.5)<br>37 (71.2) | Include<br>Exclude<br>Abstain | **16 (94.1)**<br>1 (5.9)<br>0 |
| 25 | reporting whether a published paper has open peer reviews available. |  |  | 1-3<br>4-6<br>7-9 | 9 (17.6)<br>19 (37.3)<br>23 (45.1) | Include<br>Exclude<br>Abstain | 8 (44.4)<br>10 (55.6)<br>2 |
| 26 | reporting whether a data management plan was shared |  |  | 1-3<br>4-6<br>7-9 | 11 (21.6)<br>15 (29.4)<br>25 (49.0) | Include<br>Exclude<br>Abstain | 4 (21.1)<br>15 (78.9)<br>1 |
| 27 | reporting whether data/code/materials are shared with a clear license |  |  |  |  | Include<br>Exclude<br>Abstain | **14 (87.5)**<br>2 (12.5)<br>1 |
| 28 | reporting whether the data/code/materials license is open or not |  |  | 1-3<br>4-6<br>7-9 | 6 (12.0)<br>15<br>(30.0)<br>29<br>(58.0) | Include<br>Exclude<br>Abstain | **13 (81.3)**<br>3 (18.8)<br>1 |
| 29 | reporting the use of non-proprietary software when sharing data/code/materials |  |  | 1-3<br>4-6<br>7-9 | 11<br>(21.6)<br>19<br>(37.3)<br>21<br>(41.2) | Include<br>Exclude<br>Abstain | 6 (54.5)<br>5 (45.5)<br>6 |
| 30 | reporting the use of persistent identifiers when sharing data/code/materials |  |  | 1-3<br>4-6<br>7-9 | 5 (9.6)<br>14<br>(26.9)<br>33<br>(63.5) | Include<br>Exclude<br>Abstain | **15 (88.2)**<br>2 (11.8)<br>0 |
| 31 | reporting whether workflows in computational environments were shared |  |  | 1-3<br>4-6<br>7-9 | 8 (15.4)<br>25<br>(48.1)<br>19<br>(36.5) | Include<br>Exclude<br>Abstain | 2 (13.3)<br>**13 (86.7)**<br>5 |
| 32 | reporting the (presumed) gender ratio of the authorship team |  |  | 1-3<br>4-6<br>7-9 | 23<br>(44.2)<br>21<br>(40.4)<br>8 (15.4) | Include<br>Exclude<br>Abstain | 4 (22.2)<br>14 (77.8)<br>2 |
| 33 | reporting trial results in a manuscript-style publication (peer reviewed or preprint) |  |  | 1-3<br>4-6<br>7-9 | 11<br>(21.2)<br>19<br>(36.5)<br>22<br>(42.3) | Include<br>Exclude<br>Abstain | **12 (100)**<br>0<br>5 |
| 34 | reporting systematic review results in a manuscript-style publication (peer reviewed or preprint) |  |  |  |  | Include<br>Exclude<br>Abstain | **11 (100)**<br>0<br>6 |

Participants suggested 10 novel potential core open science practices to include in Round 2 for voting, they were: use of Research Resource Identifiers (#RRID) where relevant biological resources are used in a study; inclusion of funder statements; information on whether a published paper has open peer reviews available; sharing a data management plan; use of open licenses when sharing data/code/materials; use of non-proprietary software when sharing data/code/materials; use of persistent identifiers when sharing data/code/materials; sharing research workflows in computational environments; reporting on the gender composition of the authorship team; and, reporting results of trials in a manuscript-style publication (peer-reviewed or preprint) within 2-years of study completion.

### Round 2

#### Participants

Fifty-six (70% of Round 1) participants completed the Round 2 survey (see Table 1). Of the 20 research institutions that completed Round 1, 19 (95%) institutions continued their contributions in Round 2, with up to 5 participants from each organization responding to our survey.

Researchers (N=23, 41.1%) and Research Administrators (N=11, 19.6%) again comprised most of the sample as in Round 1.

#### Round 2 voting

Of the 15 potential core open science practices that participants had not reached consensus in round 1, 6 reached consensus in Round 2. Participants agreed that the following practices were essential to reporting in the dashboard: 1) whether data were shared openly at the time of publication (with limited exceptions), 2) whether code was shared openly at the time of publication (with limited exceptions), 3) whether reporting guideline checklists were used, 4) whether author contributions were described, 5) whether ORCID identifiers were used, and 6) whether registered clinical trials were reported in the registry within 2 years of study completion.

Participants then ranked the 10 novel potential core open science practices for the first time, these items were suggested by participants in Round 1. None of these 10 new practices reached consensus in Round 2. There were no other explicitly described practices suggested by participants in Round 2 to consider for the dashboard in Round 3.

### Round 3

#### Participants

Twenty-one participants were present on day 1 of the consensus meeting and 17 on day 2 of the meeting. Full demographics are described in Table 1, one participant on each day did not provide any demographic information.

#### Round 3 voting

There were 19 items that had not reached consensus in Round 2. After discussing each item, some were re-worded slightly, expanded into two items, or collapsed into a single item (See notes on modifications made in Table 2). Ultimately, participants voted on 22 potential open science practices in Round 3. For the complete list of items voted upon, please see Appendix 4. One of these items asked participants to vote on “reporting whether registered clinical trials were reported in the registry within 1 year of study completion”. An item describing “reporting that registered clinical trials were reported in the registry within 2 years of study completion” reached consensus in Round 2; however, several participants commented that the timeframe was inconsistent with requirements of funders that have signed the World Health Organization joint statement on public disclosure of results from clinical trials, which specified 12 months. Based on this, participants were asked to re-vote on this item using the 1-year cutoff.

Of the 22 potential items voted on in Round 3, 12 reached consensus for inclusion; they were: 1) whether systematic reviews have been registered, 2) whether there was a statement about study materials sharing with publications, 3) the use of persistent identifiers when sharing data/code/materials, 4) whether data/code/materials are shared with a clear license, 5) whether the data/code/materials license is open or not, 6) citations to data, 7) what proportion of articles are published open access with a breakdown of time delay, 8) the number of preprints, 9) that registered clinical trials were reported in the registry within 1 year of study completion, 10) trial results in a manuscript-style publication (peer reviewed or preprint), 11) systematic review results in a manuscript-style publication (peer reviewed or preprint), 12) whether research articles include funding statements. One item reached consensus for exclusion from the dashboard, it was “reporting whether workflows in computational environments were shared”.

Participants agreed this item should be a component of the existing item “reporting whether code was shared openly at the time of publication (with limited exceptions)”.

The final list of the 19 items that reached consensus for inclusion is provided in Table 3. Participants discussed how some of the items that reached consensus for inclusion represented essential practices more broadly related to transparency or reporting than practices generally considered traditional open science procedures. Following Round 3 items that reached consensus were grouped based on these broad categories (traditional open science vs broader transparency practices for reporting) and participants were asked to rank order the practices based on how they should be prioritized for programming for inclusion in our proposed dashboard, these results are provided in Table 4. Items with higher scores represent those that were prioritized more. The top two traditional open science practices prioritized were: 1) reporting whether clinical trials were registered before they started recruitment; and 2) reporting whether study data were shared openly at the time of publication (with limited exceptions). The top two broader transparency practices prioritized were: 1) reporting whether author contributions were described; and 2) reporting whether author conflicts of interest were described.

**Table 3.** Items that reached consensus for inclusion in the open science dashboard

| # | Item | Round where consensus was reached |
| --- | --- | --- |
| 1 | Reporting whether clinical trials were registered before they started recruitment | 1 |
| 2 | Reporting whether study data was shared openly at the time of publication (with limited exceptions) | 1 |
| 3 | Reporting whether study code was shared openly at the time of publication (with limited exceptions) | 2 |
| 4 | Reporting whether study reporting guideline checklists were used | 2 |
| 5 | Reporting whether author conflicts of interest were reported | 2 |
| 6 | Reporting whether author contributions were reported | 2 |
| 7 | Reporting whether ORCID identifiers were reported | 2 |
| 8 | Reporting whether systematic reviews have been registered | 3 |
| 9 | Reporting whether there was a statement about study materials sharing with publications | 3 |
| 10 | Reporting the use of persistent identifiers when sharing data/code/materials | 3 |
| 11 | Reporting whether data/code/materials are shared with a clear license | 3 |
| 12 | Reporting whether the data/code/materials license is open or not | 3 |
| 13 | Reporting citations to data | 3 |
| 14 | Reporting what proportion of articles are published open access with a breakdown of time delay | 3 |
| 15 | Reporting the number of preprints | 3 |
| 16 | Reporting that registered clinical trials were reported in the registry within 1 year of study completion | 3 |
| 17 | Reporting trial results in a manuscript-style publication (peer reviewed or preprint) | 3 |
| 18 | Reporting systematic review results in a manuscript-style publication (peer reviewed or preprint) | 3 |
| 19 | Reporting whether research articles include funding statements | 3 |

**Table 4.**
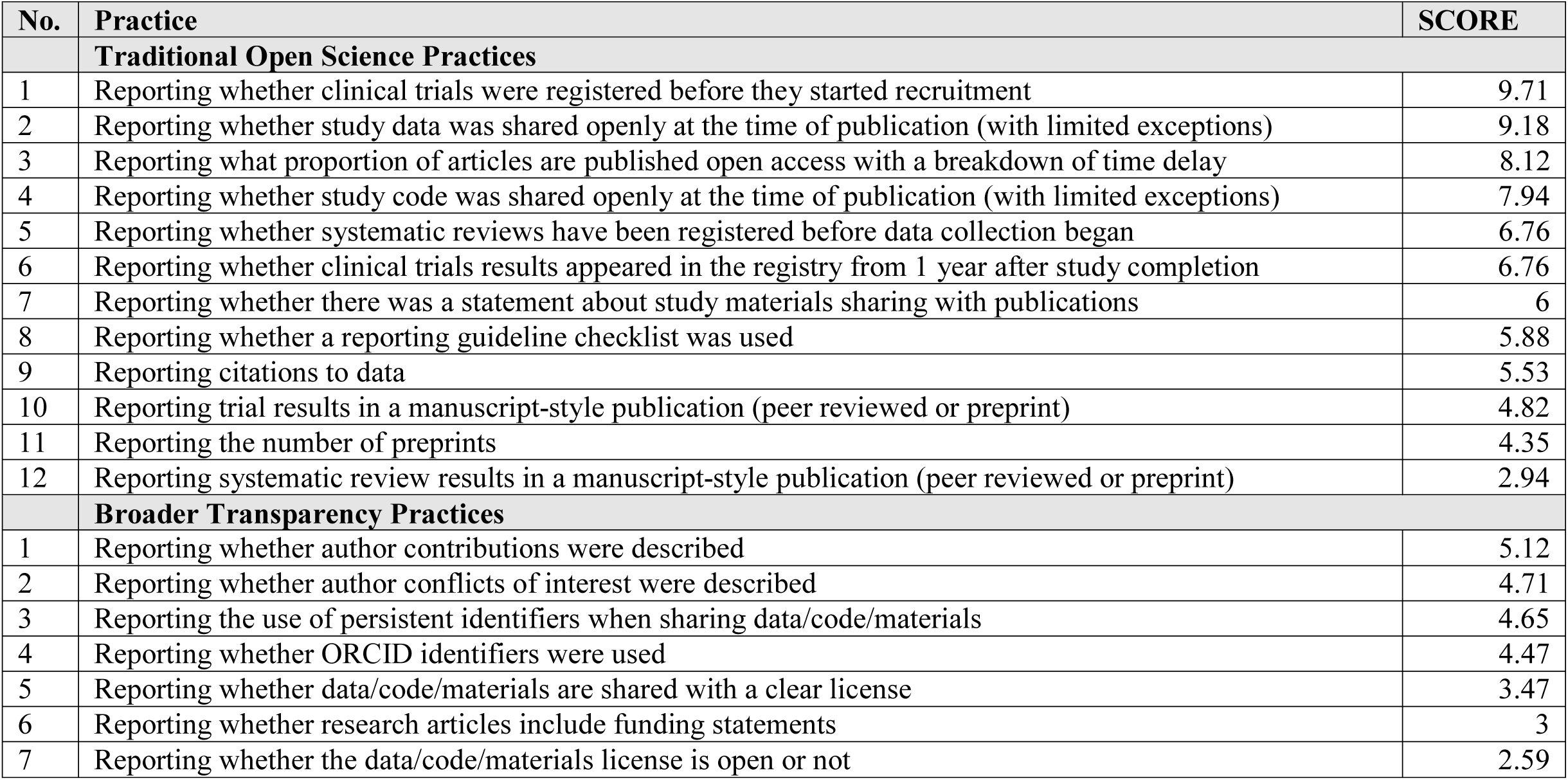
Prioritization of Traditional Open Science Practices and Broader Transparency Practices

## Discussion

This research aimed to establish a core set of open science practices to monitor at biomedical institutions. This core set of practices was developed to inform items to track in a proposed automated open science dashboard, which could be deployed by institutions and report aggregated institutional level information on performance for each practice included. The value of a consensus list of open science practices may be of broader value when developing policy, education, and interventions to improve open science in biomedicine.

Through consulting with 80 stakeholders from 20 institutions, consensus was reached on the value of tracking 19 practices in the proposed dashboard. By taking the approach of consulting the community and building consensus on the practices to include in the dashboard, we intend to develop a dashboard that best meets the needs of the community. By bringing the community together prior to developing the tool, we have also had the opportunity to brainstorm and discuss implementation strategies. We now have a roadmap to guide how to obtain community feedback on a prototype of the dashboard and a plan to pilot implementation at 6 institutions. This pilot and implementation exercise will position us to better understand barriers and enablers to adoption and use of the proposed open science dashboard^23^.

We anticipate that the open science dashboard will serve as a tool for institutions to track their progress adopting the agreed open science practices, but also to assess their performance relevant to existing mandates. For example, the dashboard will enable institutions to monitor their adherence to mandates related to open access publishing, clinical trial registration and reporting, and data sharing, all of which are commonly mandated by funders globally and related stakeholders in the research ecosystem^24–26^. We also anticipate that several of the open science practices included in the dashboard will not reflect practices that are widely performed or mandated. Some items may therefore reflect aspirational practices for the community. The dashboard can be used to benchmark for improvements in these areas.

The proposed dashboard is a necessary precursor for providing institutional feedback on the performance of the agreed open science practices. As we pilot implementation of the dashboard, we will consider how the tool can provide tailored feedback to individual institutions, or distinct settings. The central goal of the dashboard is not to facilitate comparison between institutions (i.e., where adherence to practices can be directly compared within the dashboard across different institutions). This type of ranking is counter to our community-driven initiative that seeks to provide a tool for institutional level improvement in open science rather than to pit organizations, who often are situated quite differently, against one another. Our vision is that the tool will not develop to be punitive, competitive or a prestige indicator as this is likely to further contribute to the systematic enablement of high resource institutions. Nonetheless, a core set of agreed practices is helpful for comparative meta-research around open science.

We intend for the dashboard to be implemented at the individual institution level. Understanding a given institution’s setting, current norms, and resource circumstances will be critical to deciding how to best implement the dashboard in that environment. A key step in the program to develop the proposed dashboard will be to carefully consider the appropriateness of the dashboard being publicly available versus hosted internally by biomedical institutions. Preference is likely to vary across institutions based on their circumstances.

While the structured, anonymous, and democratic approach of the Delphi process offers many advantages to reaching consensus, this study is not without limitations. While we endeavored to attract a diverse and representative sample of institutions to contribute, ultimately it is likely that the participants and institutions that agreed to take part may not be as representative of the global biomedical research culture as we desired, such as having a stronger interest in or commitment to open science. Further, our Delphi surveys and consensus meetings were conducted in English only and the meeting was not conducive for attendance across all time zones. These factors will have created barriers to participation for some institutions or participants. Defining who is an ‘expert’ to provide their views in any Delphi exercise provides an inherent challenge^27^. We faced this challenge here, especially considering the diversity of open science practices and the nuances of applying these practices in distinct biomedical sub-disciplines. For example, our vision to create a single biomedical dashboard to deploy at the institutional level may mean we have missed nuances in open science practices in preclinical as compared to clinical research.

The next phase of this research program will involve developing the open science dashboard interface and its programming. While we aim to create a fully automated tool, some core open science practices that reached consensus for inclusion in the dashboard may not lend themselves to reliable, automated analysis. In these instances, we will exclude the open science practice from monitoring. We chose not to restrict the community of Delphi participants in terms of the ease of automation of what they wanted in the tool—we encouraged participants to ‘think big’.– Ultimately, some items may not be possible to include. We anticipate iterative consultation with the community as we work to develop a dashboard that best meets their needs.

Over time, we will need to monitor the dashboard itself. As open science becomes increasingly embedded in the research ecosystem, the core practices of today may differ from those of the future. As technology changes and use of persistent identifiers on research outputs evolves, so too will our abilities to automate practices more readily. We will need to monitor and stay abreast of these changes to ensure the dashboard is sustainable and relevant over time.

## Data Availability

All data produced are available online at https://osf.io/jm8wg/

## Author Contributions

Conceptualization, funding acquisition, methodology: KDC, SH, JB, UD, DLF, LGH, JP, NR, DS, JPA, RC, CN, ES, TvL, DM; Data curation, Formal analysis, Project administration, Writing — first draft: KDC; Investigation: all authors. Validation: all authors. Writing — review & editing: all authors

## Acknowledgements

We are grateful to the participants who completed the surveys in Rounds 1 and 2 and attended the in virtual consensus meeting, including: Michaela Fritz, Christine Snidal.

## Appendix 1.

List of institutions initially identified and invited to contribute

**Appendix 1.** List of institutions initially identified and invited to contribute
| **No.** | **Institution** | **Country** |
| --- | --- | --- |
| 1 | Bond University | Australia |
| 2 | Queensland University of Technology | Australia |
| 3 | Medical University Vienna | Austria |
| 4 | UZ Leuven | Belgium |
| 5 | Universidade Federal de Pelotas | Brazil |
| 6 | Ottawa Hospital Research Institute | Canada |
| 7 | University of Calgary | Canada |
| 8 | University Health Network | Canada |
| 9 | Montreal Neurological Institute | Canada |
| 10 | Douglas Research Centre | Canada |
| 11 | University of Santiago | Chile |
| 12 | Universidad del Rosario | Colombia |
| 13 | University of Oxford | England |
| 14 | King's Health Partners | England |
| 15 | UPMC-Paris 06 University | France |
| 16 | Charité Universitätsmedizin Berlin | Germany |
| 17 | University of Hong Kong | Hong Kong, China |
| 18 | Hong Kong Baptist University | Hong Kong, China |
| 19 | Pune University | India |
| 20 | Università Vita-Salute San Raffaele | Italy |
| 21 | University of Torino | Italy |
| 22 | Istituto Italiano di Tecnologia | Italy |
| 23 | National Autonomous University of Mexico | Mexico |
| 24 | Erasmus University Medical Center Amsterdam | Netherlands |
| 25 | University of Nigeria | Nigeria |
| 26 | University of Edinburgh | Scotland |
| 27 | Stellenbosch University | South Africa |
| 28 | Vall d'Hebron Institut de Recerca | Spain |
| 29 | Karolinska University Hospital | Sweden |
| 30 | University of Basel / University Hospital Basel | Switzerland |
| 31 | Stanford University | United States of America |
| 32 | University of Alabama | United States of America |

## Appendix 2. Round 1 survey

1. What is your age?
2. Which describes you best?
Choices: female, male, transgender, non-binary, other, prefer not to say
3. Do you identify as being part of an ethnic or racial minority group? Choices: yes, no, prefer not to say
4. Which organization is your primary affiliation?
5. Which describes you best?

Choices: early-career researchers (within 5 years of first permanent position); mid-career researchers (5-10 years into permanent position); senior researchers (more than 10 years into their permanent position); library or scholarly communication staff; research administrators; faculty involved in researcher assessment (e.g., hiring and tenure committee members); other, please specify

*The aim of this project is to come up with a core set of open science practices that biomedical institutions can track over time. Once we agree on what core open science practices are important in the community, we plan to develop an automated online dashboard to track these practices over time. By tracking open science practices, we will be better able to create open science policies and measure the impact of these policies and other interventions to drive improvements in open science practices*.

*There are a wide range of possible open science practices. For each of the practices below, please indicate how important you think each is to be included in our core set of open science practices. We define core open science practices as those that are essential to perform for virtually all studies.*

6. Please indicate your response on the 1-9 scale provided (1 = Strongly disagree that this is a core open science practice; 5 =Unsure if this is a core open science practice; 9 = Strongly agree that this is a core open science practice). Please provide any comments about the item or your decision that you would like to share with the other participants.

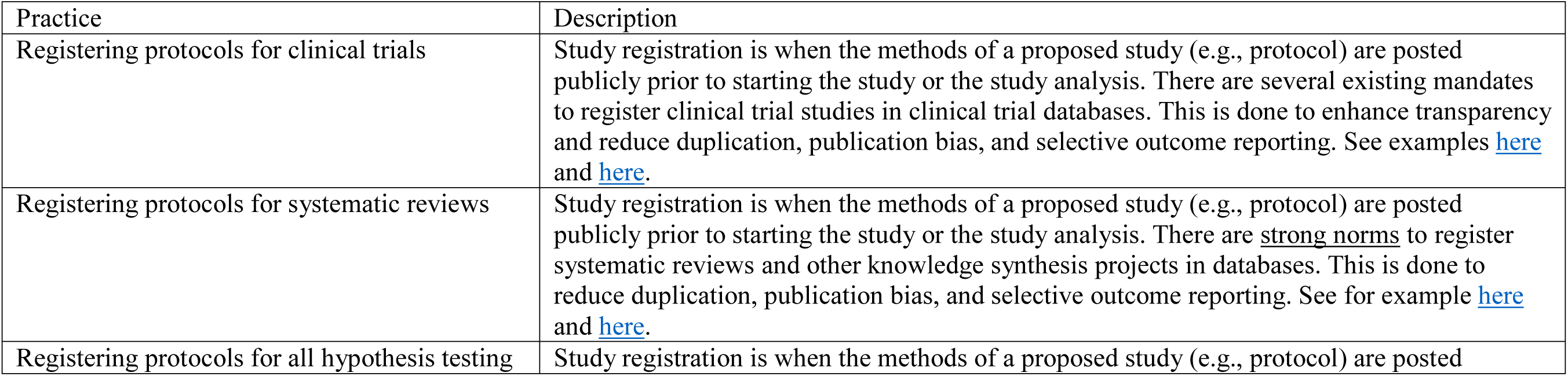
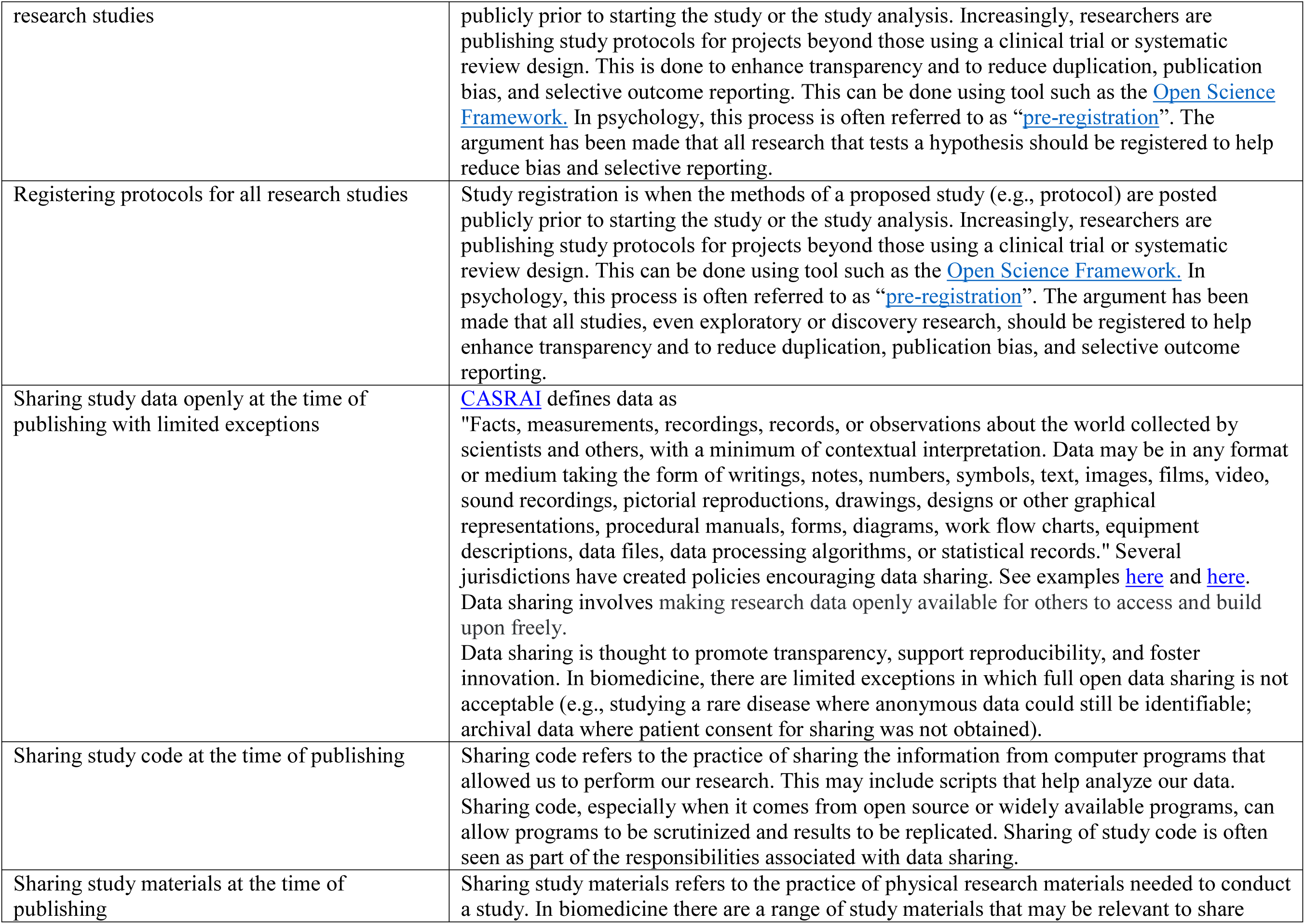
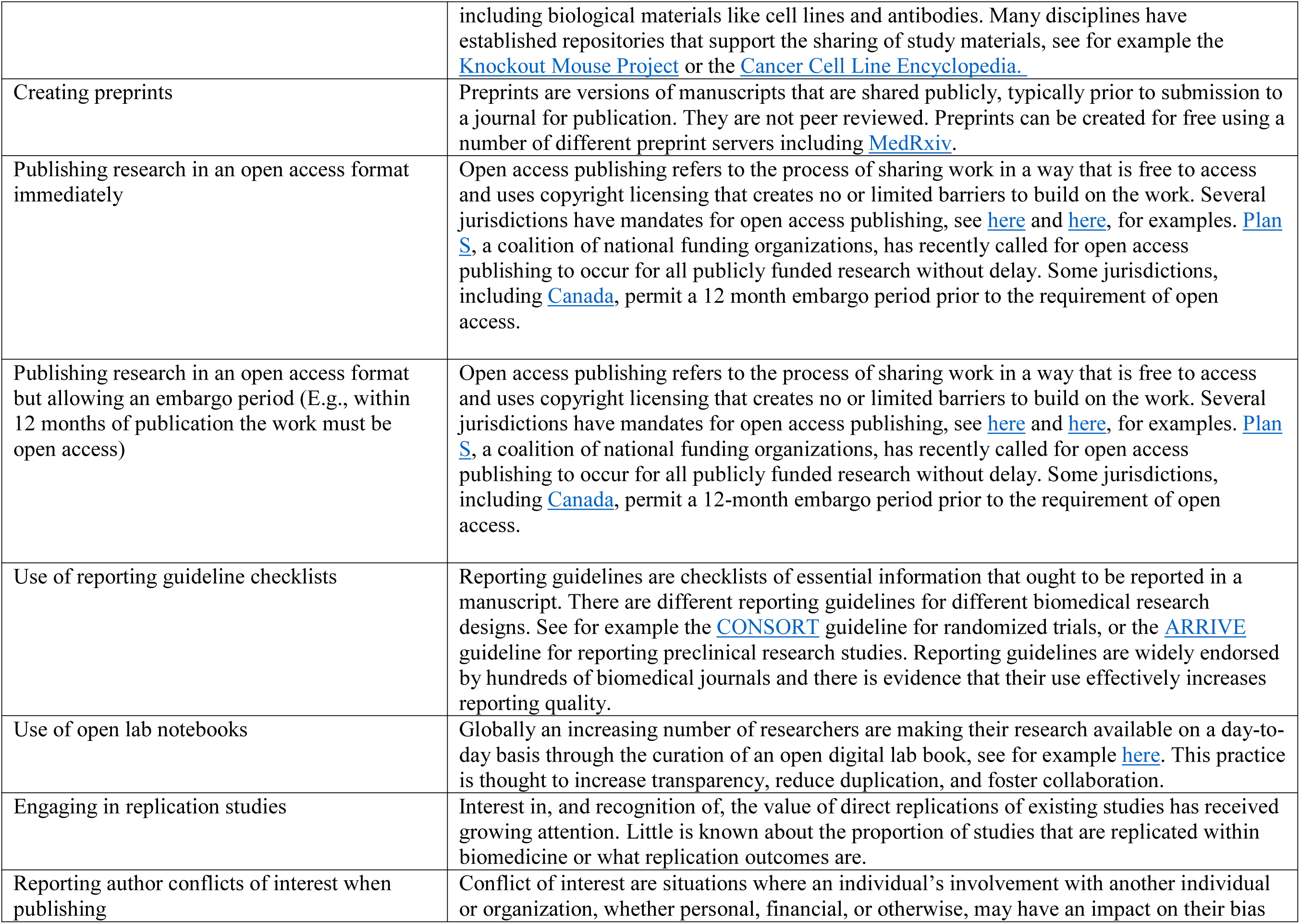
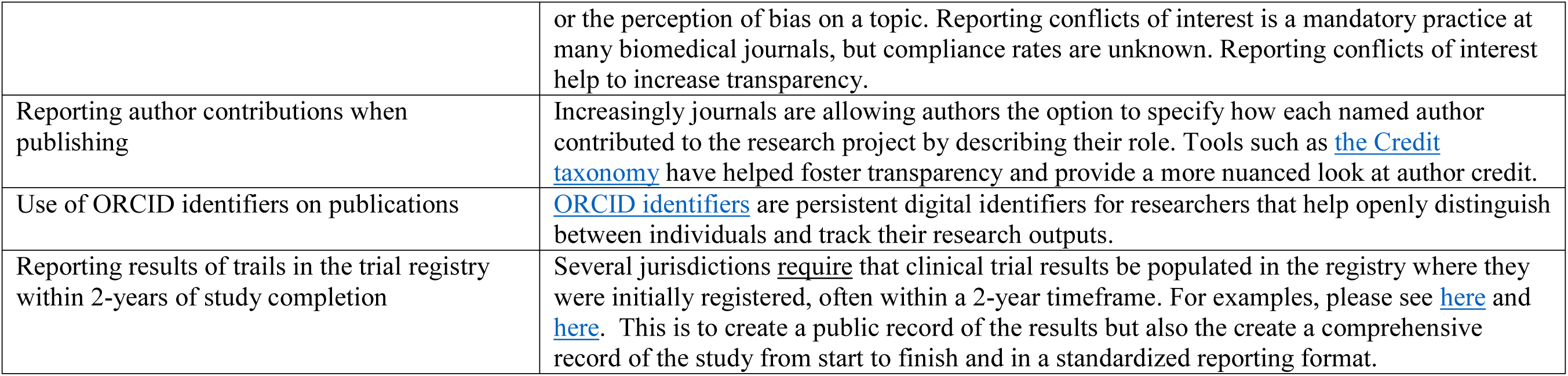

Note: for each item above participants will be presented with a comment box in which they can choose to explain their reasoning for their vote.

7. Are there any open science practices that you think are core to monitor that were not listed in the previous section? If so, please describe these here.
8. Do you have any other comments to share? If so, please describe these here.

## Appendix 3. Round 2 Survey

### Section 1

For items in this section, you are asked to use the scale:

Exclude this open science practice from monitoring
Include this open science practice for monitoring
I want to discuss this practice further in the virtual meeting as part of the 3^rd^ round of the Delphi
I don’t have enough content expertise to vote on this item

For each item you have the option of providing a comment to justify your answer.

**Note that the table of definitions provided in the Round 1 survey was again provided.**

1. Registering protocols for systematic reviews
2. Registering protocols for all hypothesis testing research studies
3. Registering protocols for all research studies
4. Sharing study data openly at the time of publishing with limited exceptions
5. Sharing study code at the time of publishing
6. Sharing study materials at the time of publishing
7. Creating preprints
8. Publishing research in an open access format immediately (ie. Gold Open Access)
9. Publishing research in an open access format but allowing an embargo period (i.e., Green Open Access)
10. Use of reporting guideline checklists
11. Use of open lab notebooks
12. Engaging in replication studies
13. Reporting author contributions when publishing
14. Use of ORCID identifiers on publications
15. Reporting results of trials in the trial registry within 2-years of study completion

### Section 2

For items in this section, you are asked to respond indicating how much you agree the item should be included in the open science dashboard, on a 1 to 9 scale, with endpoints and midpoint:

1- Strongly Disagree 5- Unsure 9- Strongly agree

For each item you have the option of providing a comment to justify your answer.

1. Use of #RRID (Research Resource Identifiers) where relevant biological resources are used in a study.
2. Resource Identifiers (#RRID) are ID numbers assigned to help researchers cite key resources (antibodies, model organisms and software projects) in the biomedical literature to improve transparency of research methods.
3. Inclusion of funder statements.
4. statements ought to specify whether the research was funded or not, if so, who the funder is and what role they had in the design and conduct of the work.
5. Information of whether a published paper has open peer reviews available.
6. pen peer review refers to the process of making peer reviewer, editorial, and author rebuttals feedback publicly available alongside a published article. Depending on the journal sometimes the names of the peer reviewer and editor are published alongside their review reports. One value of open peer review is increased transparency regarding journal operations and decision making.
7. Sharing a data management plan.
8. management plans are documents that describe how a given research project will acquire, manage, analyze, store, share and archive data resulting from the work. Some stakeholders, such as federal funders in some jurisdictions, require data management plans.
9. Use of open licenses when sharing data/code/materials.
10. data/code/materials using a CC-0, CC-BY, or similar license reduces barriers for others to access, use, and built upon shared materials. Clear use of a creative Commons license means that others don’t have to contact the authors of the data/code/materials to enquire about permissions for reuse or modification.
11. Use on non-proprietary software when sharing data/code/materials.
12. Non-proprietary software refers to software that are in the public domain that do not require licenses to access and use. Sharing data/code/materials using non-proprietary software may foster greater access.
13. Use of persistent identifiers when sharing data/code/materials.
14. identifiers are permanent references to a document. An example of a persistent identifier is a DOI. Sharing data/code/materials using persistent identifiers helps make them findable, recognizable, and more easily mapped between information systems.
15. Sharing of research workflows in computational environments.
16. biomedical research (e.g., bioinformatics) involving complex analytical pipelines, the use of workflow tools can be used to transparently present processes. E.g., Snakemake, Nextflow, Galaxy, and Apache Taverna.
17. Gender of authorship team.
18. institutions and jurisdictions are increasingly focused on equity, diversity, and inclusion in research. One aspect of this involves supporting female researchers to continue in research and in research leadership. This metric would report the overall ‘female’ contribution to the research project, e.g., the proportion of team members based on author name, that are likely to be female.
19. Reporting trial results in a manuscript within 2 years of study completion
20. result of trials in a manuscript-style publication (peer-reviewed or preprint) within 2-years of study completion.

## Appendix 4. Round 3 Survey

1. What is your age?
2. Which describes you best? (female, male, transgender, non-binary, other, prefer not to say)
3. Do you identify as being part of an ethnic or racial minority group (yes, no, prefer not to say)
4. Which organization is your primary affiliation?
5. Which describes you best? (early-career researchers (within 5 years of first permanent position); mid-career researchers (5-10 years into permanent position); senior researchers (more than 10 years into their permanent position); library or scholarly communication staff; research administrators; faculty involved in researcher assessment (e.g., hiring and tenure committee members); other, please specify.)

### Day 1

Items were voted on using the responses: include, exclude and abstain.

1. A metric reporting what proportion of articles are published open access with a breakdown of time delay
2. A metric reporting whether systematic reviews have been registered
3. A metric reporting whether hypothesis testing research has been registered
4. A metric reporting whether any research paper has been registered
5. A metric reporting whether there was a statement about study materials sharing with publications
6. A metric reporting whether open lab notebook information has been shared
7. A metric reporting citations to data
8. A metric reporting citations to code
9. A metric reporting whether a published paper has open peer reviews available
10. A metric reporting whether a data management plan has been shared in a published article
11. A metric reporting whether workflows in computational environments were shared
12. A metric reporting the (presumed) gender ratio of the authorship team

### Day 2

Items were voted on using the responses: include, exclude and abstain.

1. A metric reporting the number of preprints
2. A metric reporting whether research articles include funding statements
3. A metric reporting the use of persistent identifiers when sharing data/code/materials
4. A metric reporting the use of Research Resource Identifiers (RRID) (where relevant)
5. A metric reporting whether data/code/materials are shared with a clear license
6. A metric reporting whether the data/code/materials license is open or not
7. A metric reporting the use of non-proprietary software when sharing data/code/materials
8. A metric reporting that registered clinical trials were reported in the registry within 1 year of study completion
9. A metric reporting trial results in a manuscript-style publication (peer reviewed or preprint)
10. A metric reporting systematic review results in a manuscript-style publication (peer reviewed or preprint)

## Appendix 5. Post consensus meeting ranking survey

Below you will find two questions. The first presents what we considered to be traditional open science practice, while the second presents indicators that may be more broadly related to transparency. Please rank order the items in each list based on which you think is most relevant to prioritize for inclusion in the dashboard.

**1.** Traditional Open Science Practices: Please rank order the following items that reached consensus based on which you think is of most importance to include in the dashboard. To do so you can click and drag the items up or down.

|  |
| --- |
| A metric reporting whether clinical trials were registered before they started recruitment |
| A metric reporting whether study data was shared openly at the time of publication (with limited exceptions) |
| A metric reporting whether study code was shared openly at the time of publication (with limited exceptions) |
| A metric reporting whether study reporting guideline checklists were used |
| A metric reporting whether systematic reviews have been registered |
| A metric reporting whether there was a statement about study materials sharing with publications |
| A metric reporting citations to data |
| A metric reporting what proportion of articles are published open access with a breakdown of time delay |
| A metric reporting the number of preprints |
| A metric reporting that registered clinical trials were reported in the registry within 1 year of study completion |
| A metric reporting trial results in a manuscript-style publication (peer reviewed or preprint) |
| A metric reporting systematic review results in a manuscript-style publication (peer reviewed or preprint) |

**2.** Open science practices related to reporting transparency: Please rank order the following items that reached consensus based on which you think is of most importance to include in the dashboard. To do so you can click and drag the items up or down.2

|  |
| --- |
| A metric reporting whether author conflicts of interest were reported |
| A metric reporting whether author contributions were reported |
| A metric reporting whether ORCID identifiers were reported |
| A metric reporting the use of persistent identifiers when sharing data/code/materials |
| A metric reporting whether research articles include funding statements |
| A metric reporting whether data/code/materials are shared with a clear license |
| A metric reporting whether the data/code/materials license is open or not |

